# Salicylic acid and risk of colorectal cancer: a two sample Mendelian randomization study

**DOI:** 10.1101/2021.10.13.21262206

**Authors:** Aayah Nounu, Rebecca C Richmond, Isobel D Stewart, Praveen Surendran, Nicholas J. Wareham, Adam Butterworth, Stephanie J Weinstein, Demetrius Albanes, John A Baron, John L Hopper, Jane C Figueiredo, Polly A Newcomb, Noralane M Lindor, Graham Casey, Elizabeth A Platz, Loïc Le Marchand, Cornelia M Ulrich, Christopher I Li, Fränzel JB van Duijnhoven, Andrea Gsur, Peter T Campbell, Víctor Moreno, Pavel Vodicka, Ludmila Vodickova, Efrat Amitay, Elizabeth Alwers, Jenny Chang-Claude, Lori C Sakoda, Martha L Slattery, Robert E Schoen, Marc J Gunter, Sergi Castellví-Bel, Hyeong Rok Kim, Sun-Seog Kweon, Andrew T Chan, Li Li, Wei Zheng, D Timothy Bishop, Daniel D Buchanan, Graham G Giles, Stephen B Gruber, Gad Rennert, Zsofia K Stadler, Tabitha A Harrison, Yi Lin, Temitope O Keku, Michael O Woods, Clemens Schafmayer, Bethany Van Guelpen, Steven Gallinger, Heather Hampel, Sonja I Berndt, Paul D P Pharoah, Annika Lindblom, Alicja Wolk, Anna H Wu, Emily White, Ulrike Peters, David A Drew, Dominique Scherer, Justo Lorenzo Bermejo, Hermann Brenner, Michael Hoffmeister, Ann C Williams, Caroline L Relton

**Affiliations:** Integrative Cancer Epidemiology Programme (ICEP), Medical Research Council (MRC) Integrative Epidemiology Unit, Bristol Medical School, University of Bristol, Bristol, BS8 2BN, UK; School of Cellular and Molecular Medicine, University of Bristol, Bristol, BS8 1TD, UK; MRC Epidemiology Unit, University of Cambridge, Cambridge, United Kingdom; British Heart Foundation Cardiovascular Epidemiology Unit, Department of Public Health and Primary Care, University of Cambridge, Cambridge, UK; British Heart Foundation Centre of Research Excellence, University of Cambridge, Cambridge, UK; Health Data Research UK Cambridge, Wellcome Genome Campus and University of Cambridge, Cambridge, UK; Rutherford Fund Fellow, Department of Public Health and Primary Care, University of Cambridge, Cambridge, UK; National Institute for Health Research Blood and Transplant Research Unit in Donor Health and Genomics, University of Cambridge, Cambridge, UK; National Institute for Health Research Cambridge Biomedical Research Centre, University of Cambridge and Cambridge University Hospitals, Cambridge, UK; Division of Cancer Epidemiology and Genetics, National Cancer Institute, National Institutes of Health, Bethesda, Maryland, USA; Department of Medicine, University of North Carolina School of Medicine, Chapel Hill, North Carolina, USA; Centre for Epidemiology and Biostatistics, Melbourne School of Population and Global Health, The University of Melbourne, Melbourne, Victoria, Australia; Department of Epidemiology, School of Public Health and Institute of Health and Environment, Seoul National University, Seoul, South Korea; Department of Medicine, Samuel Oschin Comprehensive Cancer Institute, Cedars-Sinai Medical Center, Los Angeles, CA, USA; Department of Preventive Medicine, Keck School of Medicine, University of Southern California, Los Angeles, California, USA; Public Health Sciences Division, Fred Hutchinson Cancer Research Center, Seattle, Washington, USA; School of Public Health, University of Washington, Seattle, Washington, USA; Department of Health Science Research, Mayo Clinic, Scottsdale, Arizona, USA; Center for Public Health Genomics, University of Virginia, Charlottesville, Virginia, USA; Department of Epidemiology, Johns Hopkins Bloomberg School of Public Health, Baltimore, Maryland, USA; University of Hawaii Cancer Center, Honolulu, Hawaii, USA; Huntsman Cancer Institute and Department of Population Health Sciences, University of Utah, Salt Lake City, Utah, USA; Division of Human Nutrition and Health, Wageningen University & Research, Wageningen, The Netherlands; Institute of Cancer Research, Department of Medicine I, Medical University Vienna, Vienna, Austria; Department of Population Science, American Cancer Society, Atlanta, Georgia, USA; Oncology Data Analytics Program, Catalan Institute of Oncology-IDIBELL, L’Hospitalet de Llobregat, Barcelona, Spain; CIBER Epidemiología y Salud Pública (CIBERESP), Madrid, Spain; Department of Clinical Sciences, Faculty of Medicine, University of Barcelona, Barcelona, Spain; ONCOBEL Program, Bellvitge Biomedical Research Institute (IDIBELL), L’Hospitalet de Llobregat, Barcelona, Spain; Department of Molecular Biology of Cancer, Institute of Experimental Medicine of the Czech Academy of Sciences, Prague, Czech Republic; Institute of Biology and Medical Genetics, First Faculty of Medicine, Charles University, Prague, Czech Republic; Faculty of Medicine and Biomedical Center in Pilsen, Charles University, Pilsen, Czech Republic; Division of Clinical Epidemiology and Aging Research, German Cancer Research Center (DKFZ), Heidelberg, Germany; Division of Cancer Epidemiology, German Cancer Research Center (DKFZ), Heidelberg, Germany; University Medical Centre Hamburg-Eppendorf, University Cancer Centre Hamburg (UCCH), Hamburg, Germany; Division of Research, Kaiser Permanente Northern California, Oakland, California, USA; Department of Internal Medicine, University of Utah, Salt Lake City, Utah, USA; Department of Medicine and Epidemiology, University of Pittsburgh Medical Center, Pittsburgh, Pennsylvania, USA; Nutrition and Metabolism Section, International Agency for Research on Cancer, World Health Organization, Lyon, France; Gastroenterology Department, Hospital Clínic, Institut d’Investigacions Biomèdiques August Pi i Sunyer (IDIBAPS), Centro de Investigación Biomédica en Red de Enfermedades Hepáticas y Digestivas (CIBEREHD), University of Barcelona, Barcelona, Spain; Department of Surgery, Chonnam National University Hwasun Hospital and Medical School, Hwasun, Korea; Department of Preventive Medicine, Chonnam National University Medical School, Gwangju, Korea; Jeonnam Regional Cancer Center, Chonnam National University Hwasun Hospital, Hwasun, Korea; Division of Gastroenterology, Massachusetts General Hospital and Harvard Medical School, Boston, Massachusetts, USA; Channing Division of Network Medicine, Brigham and Women’s Hospital and Harvard Medical School, Boston, Massachusetts, USA; Clinical and Translational Epidemiology Unit, Massachusetts General Hospital and Harvard Medical School, Boston, Massachusetts, USA; Broad Institute of Harvard and MIT, Cambridge, Massachusetts, USA; Department of Epidemiology, Harvard T.H. Chan School of Public Health, Harvard University, Boston, Massachusetts, USA; Department of Immunology and Infectious Diseases, Harvard T.H. Chan School of Public Health, Harvard University, Boston, Massachusetts, USA; Department of Family Medicine, University of Virginia, Charlottesville, Virginia, USA; Division of Epidemiology, Department of Medicine, Vanderbilt-Ingram Cancer Center, Vanderbilt Epidemiology Center, Vanderbilt University School of Medicine, Nashville, Tennessee, USA; Leeds Institute of Cancer and Pathology, University of Leeds, Leeds, UK; Colorectal Oncogenomics Group, Department of Clinical Pathology, The University of Melbourne, Parkville, Victoria 3010 Australia; University of Melbourne Centre for Cancer Research, Victorian Comprehensive Cancer Centre, Parkville, Victoria 3010 Australia; Genetic Medicine and Family Cancer Clinic, The Royal Melbourne Hospital, Parkville, Victoria, Australia; Cancer Epidemiology Division, Cancer Council Victoria, Melbourne, Victoria, Australia; Precision Medicine, School of Clinical Sciences at Monash Health, Monash University, Clayton, Victoria, Australia; Department of Preventive Medicine & USC Norris Comprehensive Cancer Center, Keck School of Medicine, University of Southern California, Los Angeles, California, USA; Department of Community Medicine and Epidemiology, Lady Davis Carmel Medical Center, Haifa, Israel; Ruth and Bruce Rappaport Faculty of Medicine, Technion-Israel Institute of Technology, Haifa, Israel; Clalit National Cancer Control Center, Haifa, Israel; Department of Medicine, Memorial Sloan Kettering Cancer Center, New York, New York, USA; Center for Gastrointestinal Biology and Disease, University of North Carolina, Chapel Hill, North Carolina, USA; Memorial University of Newfoundland, Discipline of Genetics, St. John’s, Canada; Department of General Surgery, University Hospital Rostock, Rostock, Germany; Department of Radiation Sciences, Oncology Unit, Umeå University, Umeå, Sweden; Wallenberg Centre for Molecular Medicine, Umeå University, Umeå, Sweden; Lunenfeld Tanenbaum Research Institute, Mount Sinai Hospital, University of Toronto, Toronto, Ontario, Canada; Division of Human Genetics, Department of Internal Medicine, The Ohio State University Comprehensive Cancer Center, Columbus, Ohio, USA; Department of Public Health and Primary Care, University of Cambridge, Cambridge, UK; Department of Clinical Genetics, Karolinska University Hospital, Stockholm, Sweden; Department of Molecular Medicine and Surgery, Karolinska Institutet, Stockholm, Sweden; Institute of Environmental Medicine, Karolinska Institutet, Stockholm, Sweden; Department of Surgical Sciences, Uppsala University, Uppsala, Sweden; University of Southern California, Preventative Medicine, Los Angeles, California, USA; Department of Epidemiology, University of Washington School of Public Health, Seattle, Washington, USA; Institute of Medical Biometry and Informatics, University of Heidelberg, Im Neuenheimer Feld 130.3, Heidelberg, Germany; Division of Preventive Oncology, German Cancer Research Center (DKFZ) and National Center for Tumor Diseases (NCT), Heidelberg, Germany; German Cancer Consortium (DKTK), German Cancer Research Center (DKFZ), Heidelberg, Germany

**Author notes:** Corresponding author: Aayah Nounu, Integrative Cancer Epidemiology Programme (ICEP), Medical Research Council (MRC) Integrative Epidemiology Unit, Bristol Medical School, University of Bristol, Bristol, UK.

## Abstract

**Background:** Salicylic acid (SA) is a metabolite that can be obtained from the diet via fruit and vegetable ingestion, of which increased consumption has observationally been shown to decrease risk of colorectal cancer (CRC). Whilst primary prevention trials of SA and CRC risk are lacking, there is strong evidence from clinical trials and prospective cohort studies that aspirin (acetylsalicylic acid) is an effective primary and secondary chemopreventative agent. Since aspirin is rapidly deacetylated to form SA, it follows that SA may have a central role for aspirin chemoprevention. Through a Mendelian randomization (MR) approach, we aimed to address whether levels of SA affected CRC risk, and whether aspirin intake as a proxy for increased SA levels was required to identify an effect.

**Methods and Findings:** A two sample MR analysis was carried out using genome-wide association study summary statistics of SA from INTERVAL and EPIC-Norfolk (N= 14,149) and CRC from Colon Cancer Family Registry (CCFR), Colorectal Cancer Transdisciplinary Study (CORECT), Genetics and Epidemiology of Colorectal Cancer (GECCO) consortia and UK Biobank (55,168 cases and 65,160 controls). The Darmkrebs: Chancen der Verhütung durch Screening (DACHS) study (4,410 cases and 3,441 controls) was used for replication and stratification of aspirin-users and non-users. Single nucleotide polymorphisms (SNPs) for SA were selected via three methods: (1) Functional SNPs that influence aspirin and SA metabolising enzymes’ activity; (2) Pathway SNPs, those that are present in the coding regions of genes involved in aspirin and SA metabolism; and (3) genome-wide significant SNPs associated with levels of circulating SA.

No association was found between the functional SNPs and SA levels, therefore they were not taken forward in an MR analysis. We identified 2 pathway SNPs (explaining 0.03% of the variance in SA levels and with an F statistic of 1.74) and 1 genome-wide independent SNP (explaining 0.05% of the variance and with an F statistic of 7.44) to proxy for SA levels. Using the pathway SNPs, an inverse variance weighted approach found no association between an SD increase in SA and CRC risk (GECCO OR:1.03, 95% CI: 0.84-1.27 and DACHS OR:1.10, 95% CI:0.58-2.07) and no association was found upon stratification between aspirin users and non-users in the DACHS study (OR:0.93, 95% CI:0.23-3.73 and OR:1.24, 95% CI:0.57-2.69, respectively). Wald ratio results using the genome-wide SNP also showed no association between an SD increase in SA and CRC risk (GECCO OR: 1.08, 95% CI:0.86-1.34 and DACHS OR: 1.01, 95% CI:0.44-2.31) and no effect was observed upon stratification by aspirin use (users OR:0.66, 95% CI: 0.11-4.12 and non-users OR: 1.12, 95% CI: 0.42-2.97).

**Conclusions:** We found no evidence to suggest that an SD increase in genetically predicted SA protects against CRC risk in the general population and upon stratification by aspirin use. However, based on the calculated variance explained by the SNPs and the F statistic, we acknowledge the possibility of weak instrument bias and the need to find better instruments for SA levels.

## Introduction

Colorectal cancer (CRC) is the fourth most common cancer in the UK and worldwide (1,2). Although incidence rates among the over 50s have remained relatively stable, rates in younger age groups have increased in both the UK and US populations (3,4). This highlights a need to find better and complementary prevention strategies to reduce risk of cancer.

Salicylic acid (SA) is a dietary metabolite that can be found in various fruits, vegetables, herbs and spices (5–7). Results from a meta-analysis of 19 cohort studies found that combined intake of fruits and vegetables reduced the risk of colorectal cancer (summary Relative Risk (RR): 0.90, 95% CI: 0.83-0.98) (8). Whilst the exact components that elicit this protective effect is unknown, it has been suggested that this may be due to levels of SA found to be related to consumption of fruits and vegetables (7). In addition, salicylates can be obtained through pharmacological intervention in the form of aspirin (acetylsalicylic acid), a well-known analgesic used to treat fever, inflammation and acute pain (9), which is rapidly deacetylated to form SA (10,11) (Figure 1), the active form of the aspirin metabolic pathway (12,13). Whilst SA can be obtained from the diet, the concentrations achieved (male and female median intake from diet 4.4mg/day and 3.2mg/day, respectively(6)) are much lower than through aspirin ingestion (aspirin doses ranging between 75mg-≥325mg given daily/alternate days)(14). Therefore it is unclear whether concentrations achieved from the diet alone are sufficient to protect against cancer or whether larger doses obtained through pharmacological intervention are required.

**Figure 1.**
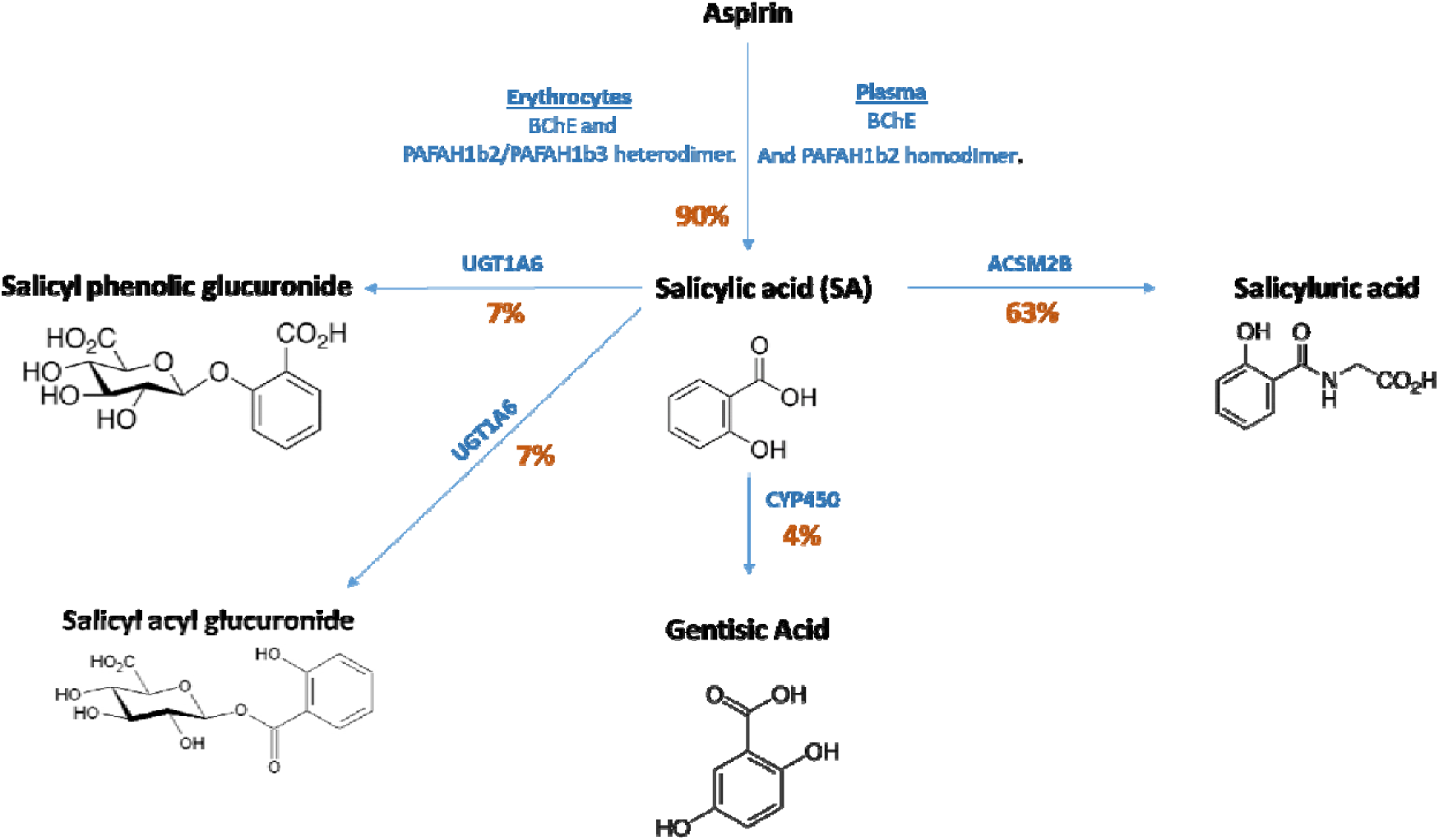
Aspirin metabolism pathway. Roughly 10% of aspirin remains unchanged and is excreted in the urine as aspirin. Aspirin is broken down into various metabolites, the most active of them being salicylic acid (13,15). Various enzymes are involved in the metabolism pathway. The percentages indicate how much of the drug is being metabolised in that pathway. Adapted from Agúndez et al (15). Abbreviations: BChE, butyrylcholinesterase; PAFAH1b2, platelet- activating factor acetylhydrolase 2; PAFAH1b3, platelet-activating factor acetylhydrolase 3; UGT1A6, UDP-glucuronosyltransferase 1-6; ACSM2B, Acyl-CoA Synthetase Medium-Chain Family Member 2B and CYP450, cytochrome P450.

As of yet, no primary prevention trials have been carried out to assess the effect of SA intervention on CRC risk, but the evidence of aspirin as a chemopreventative agent is clear (16). A long-term follow up of a randomised controlled trial (RCT) in the Women’s Health Study (WHS) showed that alternate day aspirin intake reduced the risk of CRC after a median of 17.5 years follow up (HR:0.80, 95% confidence intervals (CI):0.67-0.97) (17) and a meta-analysis of observational studies showed that aspirin is protective against CRC (relative risk (RR):0.79, 95% CI:0.74-0.85) (18). Further evidence comes from RCTs for primary and secondary prevention of vascular events. These showed that aspirin reduces the risk of CRC incidence and mortality (HR:0.76, 95% CI:0.60-0.96 and odds ratio (OR):0.79, 95% CI:0.68-0.92, respectively) (19,20). Considering aspirin is rapidly deacetylated to form SA in under 30 mins (21), and that evidence in the form of *in vivo* and *in vitro* experiments have previously shown SA to be an antiproliferative and antitumour agent(22–24), it may be that metabolism of aspirin leading to increased circulating SA levels may partially explain aspirin’s chemopreventative mode of action.

In order to identify the true effect of SA on CRC risk, conducting an RCT would be the ideal study design. However, RCTs for cancer primary prevention are lengthy and costly, therefore it would be helpful to test this association using statistical methods such as Mendelian Randomization (MR). MR uses genetic variants (mostly single nucleotide polymorphisms (SNPs)) related to modifiable factors (such as metabolite levels) to investigate the causal role of these factors on risk of disease (25–27). Through this method, MR has been likened to RCTs in that genetic variants are randomly allocated at conception the same way that an intervention is randomly allocated at the start of a trial (28,29). This lends many advantages such as overcoming the issues of confounding and reverse causation, which are commonly encountered in observational epidemiology (28). MR has previously been useful in predicting trial outcomes such as the case of selenium and prostate cancer in The Selenium and Vitamin E Cancer Prevention Trial (SELECT) (30). Results from an MR study mimicked the findings of this RCT and may have been useful to inform whether to conduct a trial that cost $114 million and that was weakly associated with increasing high-grade prostate cancer risk (31).

For this reason, we applied an MR approach using genetic “instruments” or proxies for SA to assess the causal effect of this metabolite on risk of CRC. Since aspirin is rapidly deacetylated to SA (21) and therefore a plausible proxy of increased SA levels, we also stratified our analysis between aspirin users and non-users to test the hypothesis of whether diet-derived levels of SA alone would affect risk of CRC or whether higher concentrations achieved through pharmacological intervention in the form of aspirin was required to identify an effect.

## Methods

### Genetic variants for salicylic acid

We applied a two-sample MR study design to test for the association of SA levels (sample 1) with risk of CRC (sample 2). GWAS and meta-analysis of salicylate levels were performed using 5,841 participants from the EPIC-Norfolk study (32) and 8,455 from the INTERVAL study (33). The percentage of samples with missing salicylate measurements was low (0.43% and 1.44% in EPIC-Norfolk and INTERVAL respectively), providing a total sample size of 14,149. Salicylate was measured independently in each study as one of many metabolites measured using the Metabolon DiscoveryHD4® platform (Metabolon, Inc., Durham, USA), from non-fasted plasma samples (predominantly non-fasted samples in EPIC-Norfolk) collected at baseline. Measures that were median normalised for run day were natural log transformed, winsorised to 5 standard deviations from the mean, before being regressed against age, sex and study-specific variables (measurement consignment in EPIC-Norfolk and measurement consignment, INTERVAL centre, plate number, appointment month, lag time between blood donation appointment and sample processing, and the first 5 ancestry principal components in INTERVAL) using linear regression. Residuals from this regression were standardised (mean 0, standard deviation 1) and used for further analysis. Genotyping was performed in both studies using the Affymetrix Axiom UK Biobank genotyping array. In INTERVAL, genotype imputation was performed using the combined UK10K+1000 Genomes Phase 3 reference panel. In EPIC-Norfolk, imputation was performed using the Haplotype Reference Consortium reference panel, with additional variants imputed using the UK10K+1000 Genomes Phase 3 reference panel. Genome-wide association analyses were performed using BOLT-LMM (version 2.2) (34) and variants with MAF<0.01% and INFO<0.3 were excluded. Associations from the two studies were pooled using inverse variance weighted fixed effect meta-analysis implemented in METAL (35), applying a minor allele count threshold in each study of >10.

The causal effect of SA on risk of CRC was assessed using 3 sets of genetic variants (SNPs) related to SA: (1) Functional SNPs that influence aspirin and SA metabolising enzymes’ activity (derived from Figure 1)- termed “functional SNPs”; (2) Pathway SNPs, those that are present in the coding regions of genes that are involved in aspirin and SA metabolism (based on the NCBI Build 37/UCSC hg19 from https://grch37.ensembl.org/index.html, Supplementary Table 1) termed “pathway SNPs”; (3) genome-wide significant SNPs associated with levels of circulating aspirin metabolites - termed “genome-wide SNPs”. Pathways SNPs were defined as having a Bonferroni threshold of association (P value 0.05/2701=1.85×10^−5^), an MAF≥0.01%.as well as a consistent direction of effect in both Epic-Norfolk and INTERVAL.Genome-wide signals were defined as having an association P value < 5×10^−8^ in the meta-analysis, MAF≥0.01%.consistent direction of effect across the two studies and association P value <0.01 in both studies

To account for genetic correlation, linkage disequilibrium (LD) clumping at an R^2^<0.001 and 10,000kb window was performed to retain the SNP most strongly associated with the metabolite for downstream analysis. Since an R^2^<0.001 is considered highly stringent, we also used an R^2^<0.8 to incorporate more variants while accounting for residual correlation in the model (see Statistical Analysis). An F-statistic for each SNP-exposure association was calculated to reflect the strength of the genetic instrument and indicate any possibility of weak instrument bias, usually inferred when F<10 (36). Power calculations were conducted using the mRnd online calculator to identify the OR in both directions that could be detected with the sample size available (37).

### Genetic variants for CRC incidence

SNP-outcome associations were obtained from the Colon Cancer Family Registry (CCFR), Colorectal Cancer Transdisciplinary Study (CORECT) and Genetics and Epidemiology of Colorectal Cancer (GECCO) consortia and UK Biobank (55,168 cases and 65,160 controls), hereafter collectively termed as GECCO (38–40). Genetic data from a population-based case-control study from southwestern Germany (Darmkrebs: Chancen der Verhütung durch Screening (DACHS)) was used to assess replication of the findings, and to run an MR analysis stratifyied by aspirin intake since this study has recorded aspirin use (defined as twice per week for at least a year) (41–43). This study comprised 4,410 cases of which 810 (18.37% of cases) were aspirin-users and 3340 (75.74%) were non-users, and 260 cases (5.90%) were excluded as they had reported use of other non-aspirin NSAIDs. This study also contained 3,441 controls of which 779 (22.64%) had recorded aspirin use and 2,320 (67.42%) were recorded as non-users, and 342 controls (9.94%) were excluded has they had reported use of other non-aspirin NSAIDs.

To assess the causal effect of SA on CRC risk, we tested for association in GECCO but also stratified the analysis between aspirin users and non-users in DACHS to investigate whether increased SA levels via pharmacological intervention is required to see an effect. We obtained summary association statistics from GECCO but also conducted logistic regression analyses adjusting for age and sex in the DACHS study for all the participants. We then stratified the participants of the DACHS study to aspirin users and non-users before repeating the logistic regression analyses again. Genetic instruments that had an MAF≤0.01 in both GECCO and DACHS (all participants) were excluded from further analyses.

### Statistical analyses

Analyses were carried out in R version 3.2.3 using the “Two-Sample MR” package (44). This package allows the formatting, harmonisation and analysis of summary data from genetic association studies in a semi-automated manner. The Two-Sample MR package automatically assigns effect alleles so that SNP associations with the exposure are positive i.e. so the effect allele is “metabolite-increasing”. The SNPs identified as associated with SA can then be extracted from the outcome datasets. Allele harmonization ensures that the effect (metabolite-increasing) allele in the exposure dataset is also treated as the effect allele in the outcome dataset. When only one SNP was associated with the metabolite, Wald ratios (SNP-outcome estimate ÷ SNP-exposure estimate) were calculated to assess the change in log OR per SD increase in the metabolite. When more than one SNP was available, a weighted mean weighted by the inverse variance of the Wald ratio estimates (inverse-variance weighted (IVW) method) was used to assess the causal effect of increased metabolite levels on risk of CRC incidence (45). To assess the quality of our instruments, we calculated the variance in SA levels explained by the SNPs and the F statistic. The variance explained for each SNP was calculated using the formula: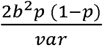, where p is the minor allele frequency, b is the SNP effect on the exposure (beta) and var is the variance of the exposure. The F statistic was calculated using the formula: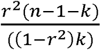 where r is the sum of the variance explained by the set of SNPs, n is the sample size of the exposure GWAS and k is the number of SNPs used to proxy the exposure. In the presence of weak instruments, we conducted an MR robust adjusted profile score (MR RAPS), which is a method that provides robust inference when many weak instruments are present (46).

Furthermore, the presence of one invalid instrument, e.g. one that is associated with exposures other than the exposure of interest (horizontal pleiotropy), may bias the results from the IVW method (47). For this reason, alternative methods that produce an unbiased estimator even when some of the genetic instruments are invalid were used as a sensitivity analysis when more than 2 SNPs were used as exposure instruments (weighted mode, weighted median and MR Egger) (44,48– 50). The MR Egger test is not constrained to pass through an effect size of 0, unlike the IVW method, allowing the assessment of the presence of directional pleiotropy through the y intercept (47,50). We also measured the Q statistic to measure the presence of pleiotropy between our instruments. If all the SNPs are valid instruments, then the individual MR estimates for each SNP will only vary by chance. A larger amount of heterogeneity would indicate that one or more of the SNPs are pleiotropic (51).

Due to the presence of a small number of independent SNPs associated with the metabolite, we also conducted a weighted generalised linear regression (WGLR) whereby SNPs in LD (R^2^<0.8) could be used with the incorporation of their correlation as weights in the regression analysis (52). This was performed using the “LDlinkR” and “MendelianRandomization” packages in R (version 3.5.1). The use of multiple SNPs explains more of the variance in the metabolite levels and therefore improves power to detect an effect (52).

## Results

### Functional SNPs and CRC risk

To interrogate the effect of SA on CRC risk, we used three methods to select our exposure instruments (Figure 2). In our first approach, we identified 4 functional SNPs that have been shown to affect enzyme efficiency in the aspirin metabolic pathway (Figure 1). For *BChE* (rs6445035), the presence of an A allele increase has been associated with a decrease in aspirin hydrolysis by around 1.2 nmol/ml/min (53). The *UGT1A6* variants rs2070959 and rs1105879 predict a higher metabolic activity of the enzyme than the wild type (54,55). Furthermore, a variant in *CYP2C9* (rs1799853) encodes an enzyme with reduced activity (56).

**Figure 2.**
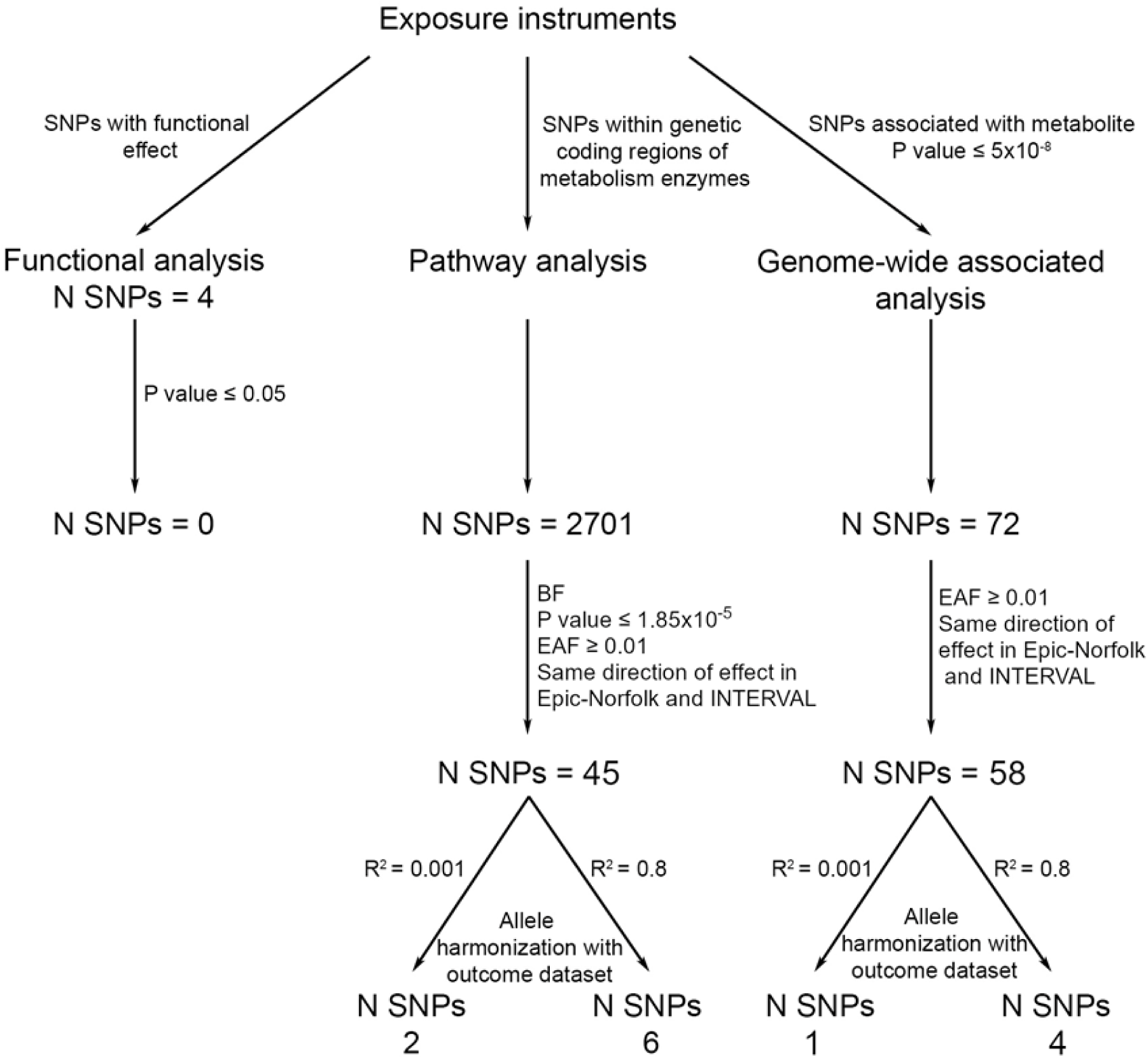
Instrument selection for functional, pathway and genome-wide SNPs. Abbreviations: SA, salicylic acid;; EAF, effect allele frequency; BF, Bonferroni.

These SNPs were tested for association with SA in the INTERVAL and (EPIC)-Norfolk study, however none of the SNPs reached nominal significance with the metabolite (Figure 3 A) (Supplementary Table 2). For this reason, these SNPs were therefore not taken forward in an MR analysis.

**Figure 3.**
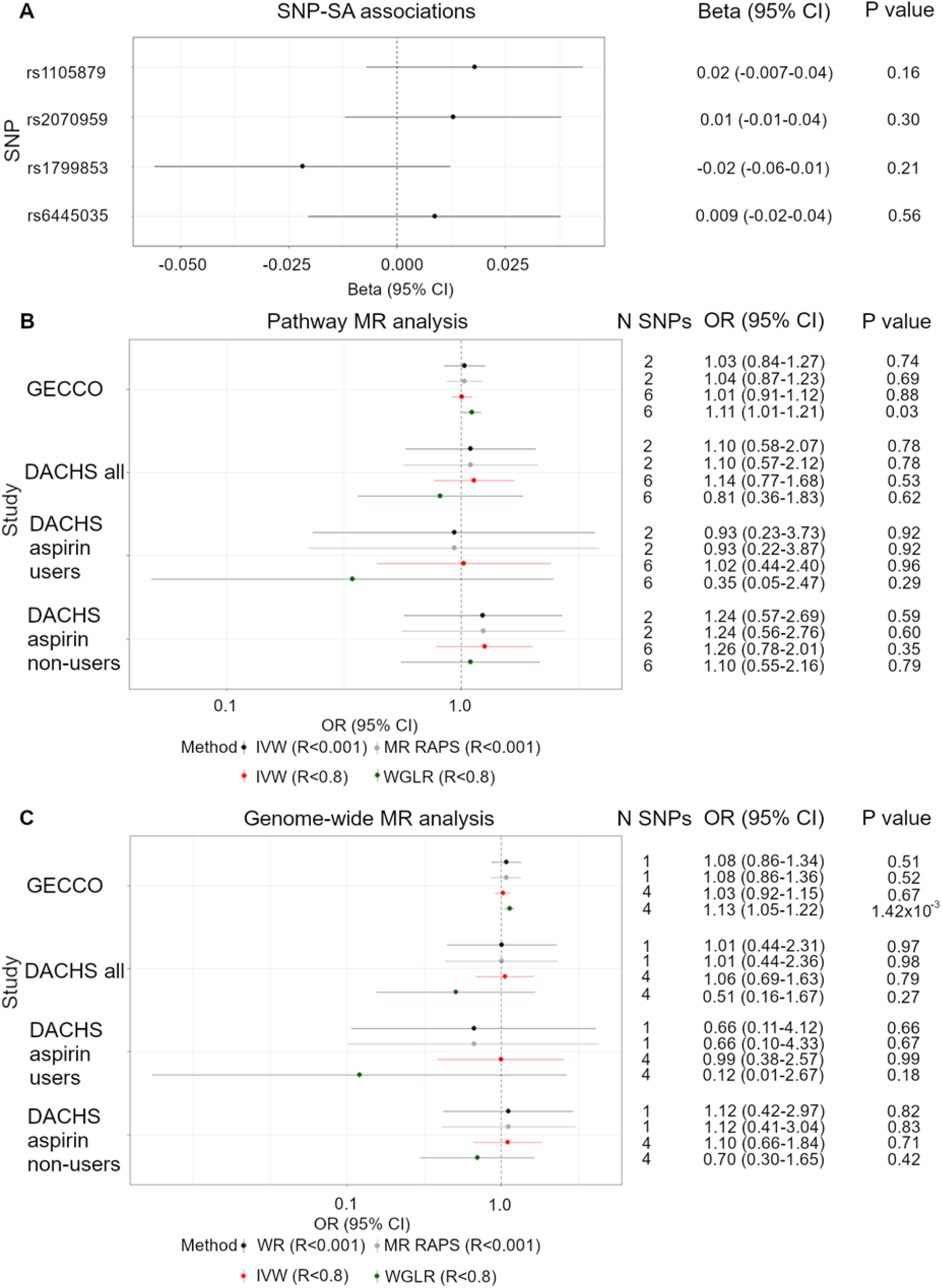
Functional SNP metabolite associations and two-sample pathway MR analysis. (A) Forest plot of single SNP associations with salicylic acid for the functional SNPs. (B) Forest plot of one SD increase in SA and its effect on CRC risk, instrumented by pathway SNPs and applying three methods: IVW after applying an LD threshold of R^2^<0.001 (black), MR RAPS after applying an LD threshold of R^2^ <0.001 (grey), IVW after applying an LD threshold of R^2^ <0.8 (red) and a WGLR after applying an LD threshold of R^2^ <0.8 (green). (C) Forest plot of one SD increase in SA and its effect on CRC risk, instrumented by genome-wide SNPs and applying three methods: WR after applying an LD threshold of R^2^ <0.001 (black), MR RAPS after applying an LD threshold of R^2^ <0.001 (grey), IVW after applying an LD threshold of R^2^ <0.8 (red) and a WGLR after applying an LD threshold of R^2^ <0.8 (green). Abbreviations: OR, odds ratio; IVW, inverse variance weighted; WGLR, weighted generalised linear regression; WR, Wald ratio; LD, linkage disequilibrium.

### Pathway SNPs and CRC risk

We investigated genetic variants within the coding regions of the enzymes involved in aspirin and SA metabolism (Figure 1). These were *BChE, PAFAH1b2, PAFAH1b3, UGT1A6, ACSM2B* and *CYP450*.

We obtained summary statistics for 2701 SNPs within the genetic coding regions of the enzymes for SA. After applying a Bonferroni threshold of association (P value 0.05/2701=1.85×10^−5^) for SNPS with consistent direction of effects in both studies and a minor allele frequency of ≥0.01 in the exposure and outcome studies, we identified 45 SNPs that could be used to instrument SA. These SNPs were then clumped at an R^2^<0.001 and 0.8, providing 2 and 6 SNPs, respectively, to instrument SA levels (Figure 2). These explained 0.03% and 0.09% of the variance in SA levels and had an F statistic of 1.74 and 2.16, respectively (Table 1).

**Table 1.** Exposure instruments used in the MR analysis

| SNP set | Study | Outcome<br>Sample size | Percentage cases<br>(%) | N SNPs | LD R<sup>2</sup> | Variance<br>explained R<sup>2</sup><br>(%) | F statistic | OR detected at 80% power |  |
| --- | --- | --- | --- | --- | --- | --- | --- | --- | --- |
|  |  |  |  |  |  |  |  | Decreased<br>risk | Increased<br>risk |
| Pathway<br>SNPs | GECCO | 120,328 | 45.85<br>55,168/120,328) | 2 | 0.001 | 0.025 | 1.74 | 0.90 | 1.11 |
|  | DACHS | 7,851 | 56.17<br>(4,410/7,851) | 2 |  |  |  | 0.68 | 1.51 |
|  | DACHS- aspirin<br>users | 1,589 | 50.98<br>(810/1,589) | 2 |  |  |  | 0.43 | 2.38 |
|  | DACHS- aspirin<br>non-users | 5,660 | 59.01<br>(3,340/5,660) | 2 |  |  |  | 0.64 | 1.64 |
|  | GECCO | 120,328 | 45.85<br>(55,168/120,328) | 6 | 0.8 | 0.092 | 2.16 | 0.95 | 1.06 |
|  | DACHS | 7,851 | 56.17<br>(4,410/7,851) | 6 |  |  |  | 0.81 | 1.24 |
|  | DACHS- aspirin<br>users | 1,589 | 50.98<br>(810/1,589) | 6 |  |  |  | 0.63 | 1.58 |
|  | DACHS- aspirin<br>non-users | 5,660 | 62.97<br>(1,165/5,660) | 6 |  |  |  | 0.78 | 1.30 |
| Genome-wide SNPs | GECCO | 120,328 | 45.85<br>(55,168/120,328) | 1 | 0.001 | 0.053 | 7.44 | 0.93 | 1.07 |
|  | DACHS | 7,851 | 56.17<br>(4,410/7,851) | 1 |  |  |  | 0.76 | 1.32 |
|  | DACHS- aspirin<br>users | 1,589 | 50.98<br>(810/1,589) | 1 |  |  |  | 0.55 | 1.83 |
|  | DACHS- aspirin<br>non-users | 5,660 | 62.97<br>(1,165/5,660) | 1 |  |  |  | 0.73 | 1.42 |
|  | GECCO | 120,328 | 45.85<br>(55,168/120,328) | 4 | 0.8 | 0.090 | 3.18 | 0.95 | 1.06 |
|  | DACHS | 7,851 | 56.17<br>(4,410/7,851) | 4 |  |  |  | 0.81 | 1.24 |
|  | DACHS- aspirin<br>users | 1,589 | 50.98<br>(810/1,589) | 4 |  |  |  | 0.63 | 1.59 |
|  | DACHS- aspirin<br>non-users | 5,660 | 62.97<br>(1,165/5,660) | 4 |  |  |  | 0.78 | 1.30 |

Abbreviations: SA, salicylic acid; LD, linkage disequilibrium; NA, not applicable; OR, odds ratio.

After LD clumping at an R^2^<0.001, 2 SNPs were taken forward in an IVW analysis but no association was found between an SD increase in SA and CRC risk (GECCO OR: 1.03, 95% CI: 0.84-1.27 and DACHS OR: 1.10, 95% CI: 0.58-2.07) (Figure 3 B). Since aspirin is rapidly deacetylated to form SA (21) and therefore a plausible proxy for increased SA levels, we stratified our analysis between aspirin users and non-users in the DACHS study. Our power calculations show that after stratification we had 80% power to detect an effect of an SD increase in SA on CRC risk with an OR of ≤0.43 and ≥2.38 in the reciprocal direction for aspirin users (N=1,589). For non-users (N=5,660), we had 80% power to detect an OR of ≤0.64 and ≥1.64 in the reciprocal direction (Table 1).However, our MR analysis showed no evidence of an association between SA and CRC risk (OR: 0.93, 95% CI: 0.23-3.73 and OR: 1.24, 95% CI: 0.57-2.69, respectively) (Figure 3 B).

The variance explained by these 2 instruments and their F statistic indicate the possibility of weak instrument bias. For this reason, we conducted MR RAPS, a method that provides robust inference even in the presence of weak instruments (46). Through this method, no association was found between an SD increase in SA and CRC risk (GECCO OR: 1.04, 95% CI: 0.87-1.23 and DACHS OR:1.10, 95% CI: 0.57-2.12), even when stratified between aspirin users and non-users (OR: 0.93, 95% CI: 0.22-3.87 and OR: 1.24, 95% CI: 0.56-2.76).

Since this LD threshold is known to be very stringent, we used a more relaxed threshold (R^2^< 0.8) to increase the number of SNPs available to instrument the metabolite and therefore explain more of the variance in SA levels. This provided 6 SNPs associated with SA (Supplementary Table 3) which showed no association between SA and CRC risk (GECCO OR: 1.01, 95% CI: 0.91-1.12 and DACHS OR:1.14, 95% CI: 0.77-1.68). Stratification between aspirin use and non-use found no association between the metabolite and CRC risk in aspirin users or non-users (OR: 1.02, 95% CI: 0.44-2.40 and OR: 1.26, 95% CI: 0.78-2.01, respectively).

Using the alternative MR methods (weighted mode, weighted median and MR Egger), no other association between SA and CRC in both GECCO and DACHS was observed, regardless of stratification (Supplementary Table 4).

Since all the SNPs were found to be on chromosome 16 (Supplementary Table 3), a WGLR method was carried out to account for the SNP correlations and include them as weights into the regression. Through this method, there was no association between SA and CRC risk in DACHS (OR:0.81, 95% CI:0.36-1.83) but a positive association in the GECCO sample (OR: 1.11, 95% CI: 1.01-1.21). No association was observed between SA and CRC risk in aspirin users or non-users (OR: 0.35, 95% CI: 0.05-2.47 and OR: 1.10, 95% CI: 0.55-2.16, respectively) (Figure 3 B). As a sensitivity analysis, the heterogeneity of the results was appraised through a Q statistic but no evidence of pleiotropy was observed-i.e. no evidence that the instruments may also be associated with another phenotype (Supplementary Table 5).

### Genome-wide significant SNPs and CRC risk

Initially, 72 SNPs were associated with SA at genome-wide significance. After applying an MAF threshold of ≥ 0.01 in the exposure and outcome studies for SNPs with a consistent direction of effect in both studies, 58 SNPs were available to instrument SA. After removing SNPs in LD at an R^2^<0.001 and R^2^<0.8, 1 SNP and 4 SNPs were available to instrument SA, respectively (Figure 2). These explained 0.05% and 0.09% of the variance in SA levels and had an F statistic of 7.44 and 3.18, respectively (Table 1).

Using the 1 independent SNP associated with SA at genome-wide significance, WR results showed no association between the genetically predicted metabolite levels and cancer risk (GECCO OR: 1.08, 95% CI: 0.86-1.34 and DACHS OR: 1.01, 95% CI:0.44-2.31). Our power calculations show that after stratification between aspirin users and non-users in the DACHS study, we had 80% power to detect an effect of an SD increase in SA on CRC risk with an OR of ≤0.55 and ≥1.83 in the reciprocal direction for aspirin users (N= 1,589). For non-users (N=5,660), we had 80% power to detect an OR of ≤0.73 and ≥1.42 in the reciprocal direction (Table 1), however, we found no association between SA levels and CRC in aspirin users (users OR: 0.66, 95% CI: 0.11-4.12 and non-users OR: 1.12, 95% CI: 0.42-2.97) (Figure 3 C).

Due to the possibility of weak instrument bias, we also conducted an MR RAPS approach, but results remained unchanged (GECCO OR: 1.08, 95% CI: 0.86-1.36, DACHS OR: 1.01, 95% CI: 0.44-2.36, DACHS aspirin users OR: 0.66, 95% CI: 0.10-4.33 and DACHS aspirin non-users OR: 1.12, 95% CI: 0.41-3.04).

To explain more of the variance, we used a less stringent LD threshold of R^2^<0.8 and therefore 4 SNPs to instrument SA (Supplementary Table 6). IVW results also showed no association between the metabolites and CRC risk (GECCO OR: 1.03, 95% CI: 0.92-1.15 and DACHS OR: 1.06, 95% CI: 0.69-1.63) and no association was found upon stratification by aspirin use (users OR: 0.99, 95% CI: 0.38-2.57, non-users OR: 1.10, 95% CI: 0.66-1.84).

Using the alternative MR methods (weighted mode, weighted median and MR Egger), no association between SA and CRC in both GECCO and DACHS was seen, regardless of stratification (Supplementary Table 7).

Since these 4 SNPs were all found on chromosome 16 (Supplementary Table 6), a WGLR method was applied to account for their correlation and found a positive association between SA and CRC risk in the GECCO sample (OR:1.13, 95% CI:1.05-1.22) but no association in the DACHS sample (OR: 0.51, 95% CI: 0.16-1.67), DACHS aspirin users (OR: 0.12, 95% CI:0.01-2.67) and DACHS aspirin non-users (OR: 0.70, 95% CI: 0.30-1.65)(Figure 3 C). As a sensitivity analysis, the heterogeneity of the results was assessed through a Q statistic but no evidence of heterogeneity was seen (Supplementary Table 8).

## Discussion

In this study, we aimed to assess whether increasing levels of SA affected risk of CRC, using an MR approach, and whether higher levels of SA proxied by pharmacological intervention in the form of aspirin use was required to identify an effect. Our analysis focused on aspirin since 90% of the drug is rapidly deacetylated to form SA (15), which is the active metabolite of the drug (12,13), and therefore increases SA levels more than would be achieved through the diet. Three different approaches were applied to identify genetic variants (instrument variables) which could serve as proxies for SA and understand the causal nature of their role in determining CRC risk. The three approaches involved selecting (i) functional, (ii) pathway and (iii) genome-wide SNPs each associated with SA. The functional genetic variants were selected through the established role of the genes in aspirin metabolism from various sources of evidence. With regards to the pathway and genome-wide significant SNPs, all were found on chromosome 16, either within or near the coding region for the enzyme *ACSM2B* which is the enzyme involved in breaking down SA into its metabolite salicyluric acid, thereby providing a plausible biological link between these SNPs and levels of SA.

We found no association between the functional SNPs and levels of SA, therefore did not take them forward to instrument SA levels. Using pathway and genome-wide SNPs, we identified 2 and 1 independent SNPs (R^2^<0.001) to proxy for SA levels, respectively, and found no association between increasing metabolite levels and CRC risk using an IVW and MR RAPS approach, regardless of aspirin stratification. Furthermore, due to the small number of instruments, we applied a less stringent LD threshold (R^2^<0.8) and identified 6 pathway SNPs and 4 genome-wide SNPs to proxy for an SD increase in SA levels. Using these SNPs, we found consistent null results using the IVW method and alternative MR methods (weighted median, weighted mode and MR Egger). However, after accounting for SNP correlation using a WGLR method, we found that an an SD increase in SA increased the risk of CRC in GECCO (OR:1.11, 95% CI:1.01-1.21, P-value:0.03 and OR:1.13, 95% CI: 1.05-1.22, P-value: 1.42×10^−3^, respectively). Overall, we found little evidence to suggest that SA affects risk of CRC, regardless of stratification.

Whilst we found no association between functional SNPs known to affect aspirin metabolism enzymes’ activity and levels of SA, this may be due to a more complex relationship between genotype and metabolite levels, rather than the assumed linear additive model. For example, with regards to the functional SNPs, Nagar et al. (2004) identified that whilst individuals with homozygous mutant alleles of *UGT1A6* had the highest metabolic activity, those that were heterozygous for alleles in 3 SNPs (including rs1105879 and rs2070959) were actually less active than homozygous wildtype enzymes(55), indicating a non-linear association between the alleles and the metabolites which is a common assumption made in regression analyses(57). This non-linear association between alleles and enzyme activity needs to also be addressed between alleles and metabolite levels to derive instrumental variables.

In order to improve the results and conclusions observed in this study, ideally we would need to identify the SNP associations with SA levels stratified between aspirin users and non-users, similar to what was carried out in our CRC outcome sample. However, to our knowledge, metabolite, genotype and phenotype data (of aspirin use) are not currently large enough to run this analysis. If a stronger association exists between the SNPs and SA levels in aspirin users, this would provide more strength of the appropriateness of the genetic instruments used to proxy for SA levels.

We also acknowledge another limitation in this study that the the measurement of metabolites was through an untargeted metabolomics approach and so the variables generated are assessed in units of measurement called “ion counts” which are calculated from the area under the curve of the corresponding peak in the mass spectrum. This means that metabolite measurements are quantitative values of relative changes as opposed to the absolute quantification of metabolite concentrations that can be achieved through targeted metabolomics (58). For this reason, it is important to focus on the direction of effect and strength of association (P-values) in this study as opposed to the magnitude of effect. This may have also impacted on the calculation of variance explained and the F statistics, which mostly indicate that the instrumental variables used in the MR were weak as they explain little of the variance and the F-statistic is below the conventionally applied indicative threshold of 10 (59). However, without carrying out a more targeted metabolomic approach and quantifying the exact effect of these SNPs on the metabolite levels, it is difficult to draw firm conclusions about the strength of the instruments used for MR. Furthermore, larger sample sizes of recorded aspirin use are required as currently, our study may have been underpowered to detect an effect hence explaining the null results using the IVW approach. Therefore, it would be useful to repeat this analysis in a larger sample with comprehensive data on aspirin use.

## Conclusions

Overall, the analyses presented have shown that dietary levels of SA as well as increased levels proxied by aspirin use may be insufficient at reducing risk of CRC, although based on the variance explained in SA levels by our SNPs and the F statistic, we acknowledge that the analysis needs to be repeated again with stronger instruments that proxy the metabolite levels.

## Data Availability

Data is available upon request from the studies themselves.

## Acknowledgements

ASTERISK: We are very grateful to Dr. Bruno Buecher without whom this project would not have existed. We also thank all those who agreed to participate in this study, including the patients and the healthy control persons, as well as all the physicians, technicians and students.

CCFR: The Colon CFR graciously thanks the generous contributions of their study participants, dedication of study staff, and the financial support from the U.S. National Cancer Institute, without which this important registry would not exist. The authors would like to thank the study participants and staff of the Seattle Colon Cancer Family Registry and the Hormones and Colon Cancer study (CORE Studies).

CLUE II: We thank the participants of Clue II and appreciate the continued efforts of the staff at the Johns Hopkins George W. Comstock Center for Public Health Research and Prevention in the conduct of the Clue II Cohort Study.

COLON and NQplus: the authors would like to thank the COLON and NQplus investigators at Wageningen University & Research and the involved clinicians in the participating hospitals.

CORSA: We kindly thank all those who contributed to the screening project Burgenland against CRC. Furthermore, we are grateful to Doris Mejri and Monika Hunjadi for laboratory assistance.

CPS-II: The authors thank the CPS-II participants and Study Management Group for their invaluable contributions to this research. The authors would also like to acknowledge the contribution to this study from central cancer registries supported through the Centers for Disease Control and Prevention National Program of Cancer Registries, and cancer registries supported by the National Cancer Institute Surveillance Epidemiology and End Results program.

Czech Republic CCS: We are thankful to all clinicians in major hospitals in the Czech Republic, without whom the study would not be practicable. We are also sincerely grateful to all patients participating in this study.

DACHS: We thank all participants and cooperating clinicians, and Ute Handte-Daub, Utz Benscheid, Muhabbet Celik and Ursula Eilber for excellent technical assistance.

EDRN: We acknowledge all the following contributors to the development of the resource: University of Pittsburgh School of Medicine, Department of Gastroenterology, Hepatology and Nutrition: Lynda Dzubinski; University of Pittsburgh School of Medicine, Department of Pathology: Michelle Bisceglia; and University of Pittsburgh School of Medicine, Department of Biomedical Informatics.

EPIC: Where authors are identified as personnel of the International Agency for Research on Cancer/World Health Organization, the authors alone are responsible for the views expressed in this article and they do not necessarily represent the decisions, policy or views of the International Agency for Research on Cancer/World Health Organization.

The EPIC-Norfolk study: we are grateful to all the participants who have been part of the project and to the many members of the study teams at the University of Cambridge who have enabled this research.

EPICOLON: We are sincerely grateful to all patients participating in this study who were recruited as part of the EPICOLON project. We acknowledge the Spanish National DNA Bank, Biobank of Hospital Clínic–IDIBAPS and Biobanco Vasco for the availability of the samples. The work was carried out (in part) at the Esther Koplowitz Centre, Barcelona.

Harvard cohorts (HPFS, NHS, PHS): The study protocol was approved by the institutional review boards of the Brigham and Women’s Hospital and Harvard T.H. Chan School of Public Health, and those of participating registries as required. We would like to thank the participants and staff of the HPFS, NHS and PHS for their valuable contributions as well as the following state cancer registries for their help: AL, AZ, AR, CA, CO, CT, DE, FL, GA, ID, IL, IN, IA, KY, LA, ME, MD, MA, MI, NE, NH, NJ, NY, NC, ND, OH, OK, OR, PA, RI, SC, TN, TX, VA, WA, WY. The authors assume full responsibility for analyses and interpretation of these data.

Interval: A complete list of the investigators and contributors to the INTERVAL trial is provided in reference (33). The academic coordinating centre would like to thank blood donor centre staff and blood donors for participating in the INTERVAL trial.

Kentucky: We would like to acknowledge the staff at the Kentucky Cancer Registry.

LCCS: We acknowledge the contributions of Jennifer Barrett, Robin Waxman, Gillian Smith and Emma Northwood in conducting this study.

NCCCS I & II: We would like to thank the study participants, and the NC Colorectal Cancer Study staff.

NSHDS investigators thank the Biobank Research Unit at Umeå University, the Västerbotten Intervention Programme, the Northern Sweden MONICA study and Region Västerbotten for providing data and samples and acknowledge the contribution from Biobank Sweden, supported by the Swedish Research Council (VR 2017-00650).

PLCO: The authors thank the PLCO Cancer Screening Trial screening center investigators and the staff from Information Management Services Inc and Westat Inc. Most importantly, we thank the study participants for their contributions that made this study possible.

SCCFR: The authors would like to thank the study participants and staff of the Hormones and Colon Cancer and Seattle Cancer Family Registry studies (CORE Studies).

SEARCH: We thank the SEARCH team.

SELECT: We thank the research and clinical staff at the sites that participated on SELECT study, without whom the trial would not have been successful. We are also grateful to the 35,533 dedicated men who participated in SELECT.

UK Biobank: We would like to thank the participants and researchers UK Biobank for their participation and acquisition of data.

WHI: The authors thank the WHI investigators and staff for their dedication, and the study participants for making the program possible. A full listing of WHI investigators can be found at: http://www.whi.org/researchers/Documents%20%20Write%20a%20Paper/WHI%20Investigator%20Short%20List.pdf

## Supporting information captions

Supplementary Table 1 – Enzyme genomic regions based on NCBI Build 37/UCSC hg19.

Supplementary Table 2 - Associations of the 4 functional SNPs with salicylic acid

Supplementary Table 3 - Pathway SNPs used as instruments

Supplementary Table 4 - Pathway SNP associations with CRC using the other MR methods

Supplementary Table 5 - Results of the Q statistic heterogeneity test for pathway SNPs

Supplementary Table 6 - Genome-wide SNPs used as instruments

Supplementary Table 7 - Genome-wide SNP associations with CRC using the other MR methods

Supplementary Table 8 - Results of the Q statistic heterogeneity test for genome-wide SNPs

## Financial Disclosure Statement

### Consortia funding

Genetics and Epidemiology of Colorectal Cancer Consortium (GECCO): National Cancer Institute, National Institutes of Health, U.S. Department of Health and Human Services (U01 CA164930, U01 CA137088, R01 CA059045, R21 CA191312, R01201407). Genotyping/Sequencing services were provided by the Center for Inherited Disease Research (CIDR) contract number HHSN268201700006I and HHSN268201200008I. This research was funded in part through the NIH/NCI Cancer Center Support Grant P30 CA015704. Scientific Computing Infrastructure at Fred Hutch funded by ORIP grant S10OD028685

ASTERISK: a Hospital Clinical Research Program (PHRC-BRD09/C) from the University Hospital Center of Nantes (CHU de Nantes) and supported by the Regional Council of Pays de la Loire, the Groupement des Entreprises Françaises dans la Lutte contre le Cancer (GEFLUC), the Association Anne de Bretagne Génétique and the Ligue Régionale Contre le Cancer (LRCC).

The ATBC Study is supported by the Intramural Research Program of the U.S. National Cancer Institute, National Institutes of Health.

CLUE II funding was from the National Cancer Institute (U01 CA86308, Early Detection Research Network; P30 CA006973), National Institute on Aging (U01 AG18033), and the American Institute for Cancer Research. The content of this publication does not necessarily reflect the views or policies of the Department of Health and Human Services, nor does mention of trade names, commercial products, or organizations imply endorsement by the US government.

Maryland Cancer Registry (MCR) cancer data was provided by the Maryland Cancer Registry, Center for Cancer Prevention and Control, Maryland Department of Health, with funding from the State of Maryland and the Maryland Cigarette Restitution Fund. The collection and availability of cancer registry data is also supported by the Cooperative Agreement NU58DP006333, funded by the Centers for Disease Control and Prevention. Its contents are solely the responsibility of the authors and do not necessarily represent the official views of the Centers for Disease Control and Prevention or the Department of Health and Human Services.

ColoCare: This work was supported by the National Institutes of Health (grant numbers R01 CA189184 (Li/Ulrich), U01 CA206110 (Ulrich/Li/Siegel/Figueireido/Colditz, 2P30CA015704-40 (Gilliland), R01 CA207371 (Ulrich/Li)), the Matthias Lackas-Foundation, the German Consortium for Translational Cancer Research, and the EU TRANSCAN initiative.

The Colon Cancer Family Registry (CCFR, www.coloncfr.org) is supported in part by funding from the National Cancer Institute (NCI), National Institutes of Health (NIH) (award U01 CA167551). The CCFR Set-1 (Illumina 1M/1M-Duo) and Set-2 (Illumina Omni1-Quad) scans were supported by NIH awards U01 CA122839 and R01 CA143247 (to GC). The CCFR Set-3 (Affymetrix Axiom CORECT Set array) was supported by NIH award U19 CA148107 and R01 CA81488 (to SBG). The CCFR Set-4 (Illumina OncoArray 600K SNP array) was supported by NIH award U19 CA148107 (to SBG) and by the Center for Inherited Disease Research (CIDR), which is funded by the NIH to the Johns Hopkins University, contract number HHSN268201200008I. The SCCFR Illumina HumanCytoSNP array was supported through NCI award R01 CA076366 (to PAN). Additional funding for the OFCCR/ARCTIC was through award GL201-043 from the Ontario Research Fund (to BWZ), award 112746 from the Canadian Institutes of Health Research (to TJH), through a Cancer Risk Evaluation (CaRE) Program grant from the Canadian Cancer Society (to SG), and through generous support from the Ontario Ministry of Research and Innovation. The content of this manuscript does not necessarily reflect the views or policies of the NCI, NIH or any of the collaborating centers in the Colon Cancer Family Registry (CCFR), nor does mention of trade names, commercial products, or organizations imply endorsement by the US Government, any cancer registry, or the CCFR.

COLON: The COLON study is sponsored by Wereld Kanker Onderzoek Fonds, including funds from grant 2014/1179 as part of the World Cancer Research Fund International Regular Grant Programme, by Alpe d’Huzes and the Dutch Cancer Society (UM 2012–5653, UW 2013-5927, UW2015-7946), and by TRANSCAN (JTC2012-MetaboCCC, JTC2013-FOCUS). The Nqplus study is sponsored by a ZonMW investment grant (98-10030); by PREVIEW, the project PREVention of diabetes through lifestyle intervention and population studies in Europe and around the World (PREVIEW) project which received funding from the European Union Seventh Framework Programme (FP7/2007–2013) under grant no. 312057; by funds from TI Food and Nutrition (cardiovascular health theme), a public–private partnership on precompetitive research in food and nutrition; and by FOODBALL, the Food Biomarker Alliance, a project from JPI Healthy Diet for a Healthy Life.

Colorectal Cancer Transdisciplinary (CORECT) Study: The CORECT Study was supported by the National Cancer Institute, National Institutes of Health (NCI/NIH), U.S. Department of Health and Human Services (grant numbers U19 CA148107, R01 CA81488, P30 CA014089, R01 CA197350,; P01 CA196569; R01 CA201407) and National Institutes of Environmental Health Sciences, National Institutes of Health (grant number T32 ES013678).

CORSA: “Österreichische Nationalbank Jubiläumsfondsprojekt” (12511) and Austrian Research Funding Agency (FFG) grant 829675.

CPS-II: The American Cancer Society funds the creation, maintenance, and updating of the Cancer Prevention Study-II (CPS-II) cohort. This study was conducted with Institutional Review Board approval.

CRCGEN: Colorectal Cancer Genetics & Genomics, Spanish study was supported by Instituto de Salud Carlos III, co-funded by FEDER funds –a way to build Europe– (grants PI14-613 and PI09-1286), Agency for Management of University and Research Grants (AGAUR) of the Catalan Government (grant 2017SGR723), and Junta de Castilla y León (grant LE22A10-2). Sample collection of this work was supported by the Xarxa de Bancs de Tumors de Catalunya sponsored by Pla Director d’Oncología de Catalunya (XBTC), Plataforma Biobancos PT13/0010/0013 and ICOBIOBANC, sponsored by the Catalan Institute of Oncology.

Czech Republic CCS: This work was supported by the Czech Science Foundation (20-03997S) and by the Grant Agency of the Ministry of Health of the Czech Republic (grants NV18/03/00199 and NU21-07-00247).

DACHS: This work was supported by the German Research Council (BR 1704/6-1, BR 1704/6-3, BR 1704/6-4, CH 117/1-1, HO 5117/2-1, HE 5998/2-1, KL 2354/3-1, RO 2270/8-1 and BR 1704/17-1), the

Interdisciplinary Research Program of the National Center for Tumor Diseases (NCT), Germany, and the German Federal Ministry of Education and Research (01KH0404, 01ER0814, 01ER0815, 01ER1505A and 01ER1505B).

DALS: National Institutes of Health (R01 CA48998 to M. L. Slattery).

EDRN: This work is funded and supported by the NCI, EDRN Grant (U01 CA 84968-06).

EPIC: The coordination of EPIC is financially supported by the European Commission (DGSANCO) and the International Agency for Research on Cancer. The national cohorts are supported by Danish Cancer Society (Denmark); Ligue Contre le Cancer, Institut Gustave Roussy, Mutuelle Générale de l’Education Nationale, Institut National de la Santé et de la Recherche Médicale (INSERM) (France); German Cancer Aid, German Cancer Research Center (DKFZ), Federal Ministry of Education and Research (BMBF), Deutsche Krebshilfe, Deutsches Krebsforschungszentrum and Federal Ministry of Education and Research (Germany); the Hellenic Health Foundation (Greece); Associazione Italiana per la Ricerca sul Cancro-AIRCItaly and National Research Council (Italy); Dutch Ministry of Public Health, Welfare and Sports (VWS), Netherlands Cancer Registry (NKR), LK Research Funds, Dutch Prevention Funds, Dutch ZON (Zorg Onderzoek Nederland), World Cancer Research Fund (WCRF), Statistics Netherlands (The Netherlands); ERC-2009-AdG 232997 and Nordforsk, Nordic Centre of Excellence programme on Food, Nutrition and Health (Norway); Health Research Fund (FIS), PI13/00061 to Granada, PI13/01162 to EPIC-Murcia, Regional Governments of Andalucía, Asturias, Basque Country, Murcia and Navarra, ISCIII RETIC (RD06/0020) (Spain); Swedish Cancer Society, Swedish Research Council and County Councils of Skåne and Västerbotten (Sweden); Cancer Research UK (14136 to EPIC-Norfolk; C570/A16491 and C8221/A19170 to EPIC-Oxford), Medical Research Council (1000143 to EPIC-Norfolk, MR/M012190/1 to EPICOxford) (United Kingdom).

The EPIC-Norfolk study (https://doi.org/10.22025/2019.10.105.00004) has received funding from the Medical Research Council (MR/N003284/1, MC-UU_12015/1 and MC_UU_00006/1) and Cancer Research UK (C864/A14136). The genetics work in the EPIC-Norfolk study was funded by the Medical Research Council (MC_PC_13048). Metabolite measurements in the EPIC-Norfolk study were supported by the MRC Cambridge Initiative in Metabolic Science (MR/L00002/1) and the Innovative Medicines Initiative Joint Undertaking under EMIF grant agreement no. 115372.

EPICOLON: This work was supported by grants from Fondo de Investigación Sanitaria/FEDER (PI08/0024, PI08/1276, PS09/02368, PI11/00219, PI11/00681, PI14/00173, PI14/00230, PI17/00509, 17/00878, PI20/00113, PI20/00226, Acción Transversal de Cáncer), Xunta de Galicia (PGIDIT07PXIB9101209PR), Ministerio de Economia y Competitividad (SAF07-64873, SAF 2010-19273, SAF2014-54453R), Fundación Científica de la Asociación Española contra el Cáncer (GCB13131592CAST), Beca Grupo de Trabajo “Oncología” AEG (Asociación Española de Gastroenterología), Fundación Privada Olga Torres, FP7 CHIBCHA Consortium, Agència de Gestió d’Ajuts Universitaris i de Recerca (AGAUR, Generalitat de Catalunya, 2014SGR135, 2014SGR255, 2017SGR21, 2017SGR653), Catalan Tumour Bank Network (Pla Director d’Oncologia, Generalitat de Catalunya), PERIS (SLT002/16/00398, Generalitat de Catalunya), CERCA Programme (Generalitat de Catalunya) and COST Actions BM1206 and CA17118. CIBERehd is funded by the Instituto de Salud Carlos III.

ESTHER/VERDI. This work was supported by grants from the Baden-Württemberg Ministry of Science, Research and Arts and the German Cancer Aid.

Harvard cohorts (HPFS, NHS, PHS): HPFS is supported by the National Institutes of Health (P01 CA055075, UM1 CA167552, U01 CA167552, R01 CA137178, R01 CA151993, R35 CA197735, K07 CA190673, and P50 CA127003), NHS by the National Institutes of Health (R01 CA137178, P01 CA087969, UM1 CA186107, R01 CA151993, R35 CA197735, K07CA190673, and P50 CA127003) and PHS by the National Institutes of Health (R01 CA042182). Hawaii Adenoma Study: NCI grants R01 CA72520.

HCES-CRC: the Hwasun Cancer Epidemiology Study–Colon and Rectum Cancer (HCES-CRC; grants from Chonnam National University Hwasun Hospital, HCRI15011-1).

Interval: Participants in the INTERVAL randomised controlled trial were recruited with the active collaboration of NHS Blood and Transplant England (www.nhsbt.nhs.uk), which has supported field work and other elements of the trial. DNA extraction and genotyping was co-funded by the National Institute for Health Research (NIHR), the NIHR BioResource (http://bioresource.nihr.ac.uk) and the NIHR [Cambridge Biomedical Research Centre at the Cambridge University Hospitals NHS Foundation Trust] [*]. Metabolon Metabolomics assays were funded by the NIHR BioResource and the NIHR [Cambridge Biomedical Research Centre at the Cambridge University Hospitals NHS Foundation Trust] [*]. The academic coordinating centre for INTERVAL was supported by core funding from: NIHR Blood and Transplant Research Unit in Donor Health and Genomics (NIHR BTRU-2014-10024), UK Medical Research Council (MR/L003120/1), British Heart Foundation (SP/09/002; RG/13/13/30194; RG/18/13/33946) and the NIHR [Cambridge Biomedical Research Centre at the Cambridge University Hospitals NHS Foundation Trust]. The views expressed are those of the authors and not necessarily those of the NHS, the NIHR or the Department of Health and Social Care.

This work was supported by Health Data Research UK, which is funded by the UK Medical Research Council, Engineering and Physical Sciences Research Council, Economic and Social Research Council, Department of Health and Social Care (England), Chief Scientist Office of the Scottish Government Health and Social Care Directorates, Health and Social Care Research and Development Division (Welsh Government), Public Health Agency (Northern Ireland), British Heart Foundation and Wellcome.

Kentucky: This work was supported by the following grant support: Clinical Investigator Award from Damon Runyon Cancer Research Foundation (CI-8); NCI R01CA136726.

LCCS: The Leeds Colorectal Cancer Study was funded by the Food Standards Agency and Cancer Research UK Programme Award (C588/A19167).

Melbourne Collaborative Cohort Study (MCCS) cohort recruitment was funded by VicHealth and Cancer Council Victoria. The MCCS was further augmented by Australian National Health and Medical Research Council grants 209057, 396414 and 1074383 and by infrastructure provided by Cancer Council Victoria. Cases and their vital status were ascertained through the Victorian Cancer Registry and the Australian Institute of Health and Welfare, including the National Death Index and the Australian Cancer Database.

Multiethnic Cohort (MEC) Study: National Institutes of Health (R37 CA54281, P01 CA033619, R01 CA063464 and U01 CA164973).

MECC: This work was supported by the National Institutes of Health, U.S. Department of Health and Human Services (R01 CA81488 to SBG and GR).

MSKCC: The work at Sloan Kettering in New York was supported by the Robert and Kate Niehaus Center for Inherited Cancer Genomics and the Romeo Milio Foundation. Moffitt: This work was supported by funding from the National Institutes of Health (grant numbers R01 CA189184, P30 CA076292), Florida Department of Health Bankhead-Coley Grant 09BN-13, and the University of South Florida Oehler Foundation. Moffitt contributions were supported in part by the Total Cancer Care Initiative, Collaborative Data Services Core, and Tissue Core at the H. Lee Moffitt Cancer Center & Research Institute, a National Cancer Institute-designated Comprehensive Cancer Center (grant number P30 CA076292).

NCCCS I & II: We acknowledge funding support for this project from the National Institutes of Health, R01 CA66635 and P30 DK034987.

NFCCR: This work was supported by an Interdisciplinary Health Research Team award from the Canadian Institutes of Health Research (CRT 43821); the National Institutes of Health, U.S. Department of Health and Human Serivces (U01 CA74783); and National Cancer Institute of Canada grants (18223 and 18226). The authors wish to acknowledge the contribution of Alexandre Belisle and the genotyping team of the McGill University and Génome Québec Innovation Centre, Montréal, Canada, for genotyping the Sequenom panel in the NFCCR samples. Funding was provided to Michael O. Woods by the Canadian Cancer Society Research Institute.

NSHDS: Swedish Research Council; Swedish Cancer Society; Cutting-Edge Research Grant and other grants from Region Västerbotten; Knut and Alice Wallenberg Foundation; Lion’s Cancer Research Foundation at Umeå University; the Cancer Research Foundation in Northern Sweden; and the Faculty of Medicine, Umeå University, Umeå, Sweden.

OFCCR: The Ontario Familial Colorectal Cancer Registry was supported in part by the National Cancer Institute (NCI) of the National Institutes of Health (NIH) under award U01 CA167551 and award U01/U24 CA074783 (to SG). Additional funding for the OFCCR and ARCTIC testing and genetic analysis was through and a Canadian Cancer Society CaRE (Cancer Risk Evaluation) program grant and Ontario Research Fund award GL201-043 (to BWZ), through the Canadian Institutes of Health Research award 112746 (to TJH), and through generous support from the Ontario Ministry of Research and Innovation.OSUMC: OCCPI funding was provided by Pelotonia and HNPCC funding was provided by the NCI (CA16058 and CA67941).

PLCO: Intramural Research Program of the Division of Cancer Epidemiology and Genetics and supported by contracts from the Division of Cancer Prevention, National Cancer Institute, NIH, DHHS. Funding was provided by National Institutes of Health (NIH), Genes, Environment and Health Initiative (GEI) Z01 CP 010200, NIH U01 HG004446, and NIH GEI U01 HG 004438.

SCCFR: The Seattle Colon Cancer Family Registry was supported in part by the National Cancer Institute (NCI) of the National Institutes of Health (NIH) under awards U01 CA167551, U01 CA074794 (to JDP), and awards U24 CA074794 and R01 CA076366 (to PAN).

SEARCH: The University of Cambridge has received salary support in respect of PDPP from the NHS in the East of England through the Clinical Academic Reserve. Cancer Research UK (C490/A16561); the UK National Institute for Health Research Biomedical Research Centres at the University of Cambridge.

SELECT: Research reported in this publication was supported in part by the National Cancer Institute of the National Institutes of Health under Award Numbers U10 CA37429 (CD Blanke), and UM1 CA182883 (CM Tangen/IM Thompson). The content is solely the responsibility of the authors and does not necessarily represent the official views of the National Institutes of Health.

SMS: This work was supported by the National Cancer Institute (grant P01 CA074184 to J.D.P. and P.A.N., grants R01 CA097325, R03 CA153323, and K05 CA152715 to P.A.N., and the National Center for Advancing Translational Sciences at the National Institutes of Health (grant KL2 TR000421 to A.N.B.-H.)

The Swedish Low-risk Colorectal Cancer Study: The study was supported by grants from the Swedish research council; K2015-55X-22674-01-4, K2008-55X-20157-03-3, K2006-72X-20157-01-2 and the Stockholm County Council (ALF project).

Swedish Mammography Cohort and Cohort of Swedish Men: This work is supported by the Swedish Research Council /Infrastructure grant, the Swedish Cancer Foundation, and the Karolinska Institute’s Distinguished Professor Award to Alicja Wolk.

UK Biobank: This research has been conducted using the UK Biobank Resource under Application Number 8614

VITAL: National Institutes of Health (K05 CA154337).

WHI: The WHI program is funded by the National Heart, Lung, and Blood Institute, National Institutes of Health, U.S. Department of Health and Human Services through contracts HHSN268201100046C, HHSN268201100001C, HHSN268201100002C, HHSN268201100003C, HHSN268201100004C, and HHSN271201100004C.

