## Supplementary information for "Salicylic acid and risk of colorectal cancer: a two sample Mendelian randomization study"

Supplementary Table 1- Enzyme genomic regions based on NCBI Build 37/UCSC hg19.

| Gene | Chromosome | Start site | End site |
| --- | --- | --- | --- |
| *BChE* | 3 | 165,490,692 | 165,555,260 |
| *PAFAH1B2* | 11 | 117,014,983 | 117,047,610 |
| *PAFAH1B3* | 19 | 42,297,033 | 42,303,546 |
| *UGT1A6* | 2 | 234,600,253 | 234,681,946 |
| *CYP2C9* | 10 | 96,698,415 | 96,749,147 |
| *ACSM2B* | 16 | 20,547,548 | 20,587,749 |

Supplementary Table 2- Associations of the 4 functional SNPs with salicylic acid

| SNP | Metabolite | Effect allele | Other allele | Beta | SE | LCI | UCI | EAF | Chr | Position | P value | Sample size | Gene |
| --- | --- | --- | --- | --- | --- | --- | --- | --- | --- | --- | --- | --- | --- |
| rs6445035 | Salicylic acid | A | G | 8.71x10^-3^ | 1.49x10^-2^ | -0.02 | 0.04 | 0.20 | 3 | 165480100 | 0.56 | 14149 | *BChE* |
| rs2070959 | Salicylic acid | G | A | 0.01 | 1.28x10^-2^ | -0.01 | 0.04 | 0.32 | 2 | 234602191 | 0.30 | 14149 | *UGT1A6* |
| rs1105879 | Salicylic acid | C | A | 0.02 | 1.26x10^-2^ | -6.79x10^-3^ | 0.04 | 0.34 | 2 | 234602202 | 0.16 | 14149 | *UGT1A6* |
| rs1799853 | Salicylic acid | T | C | -0.02 | 1.75x10^-2^ | -0.06 | 0.01 | 0.14 | 10 | 96702047 | 0.21 | 14149 | *CYP2C9* |

Supplementary Table 3-Pathway SNPs used as instruments

| SNP | Effect allele | Other allele | Beta | SE | LCI | UCI | EAF | Direction | Chr | Position | P value | Sample size | LD clumping R^2^ |
| --- | --- | --- | --- | --- | --- | --- | --- | --- | --- | --- | --- | --- | --- |
| rs8062555 | C | G | -0.27 | 0.04 | -0.35 | 1.74 | 0.9792 | -- | 16 | 20566287 | 2.07x10^-10^ | 14149 | 0.001 and 0.8 |
| rs9922093 | T | C | -0.18 | 0.03 | -0.24 | 1.82 | 0.9666 | -- | 16 | 20568843 | 1.01 x10^-07^ | 14149 | 0.001 and 0.8 |
| rs146980165 | A | T | 0.24 | 0.05 | 0.15 | 2.25 | 0.0168 | ++ | 16 | 20576746 | 2.42 x10^-07^ | 14149 | 0.8 |
| rs7499557 | T | G | -0.25 | 0.04 | -0.34 | 1.75 | 0.9788 | -- | 16 | 20557620 | 1.29 x10^-09^ | 14149 | 0.8 |
| rs7500194 | A | G | 0.08 | 0.02 | 0.05 | 2.06 | 0.8573 | ++ | 16 | 20549505 | 1.90x10^-06^ | 14149 | 0.8 |
| rs8056693 | T | C | 0.18 | 0.04 | 0.11 | 2.18 | 0.0298 | ++ | 16 | 20570661 | 2.94x10^-07^ | 14149 | 0.8 |

Abbreviations: SE, standard error; LCI, lower confidence interval; UCI, upper confidence interval; Chr, chromosome; LD, linkage disequilibrium.

Supplementary Table 4- Pathway SNP associations with CRC using the other MR methods

| Study | Method | N SNPs | OR | LCI | UCI | P value |
| --- | --- | --- | --- | --- | --- | --- |
| GECCO | MR Egger | 6 | 0.99 | 0.78 | 1.27 | 0.97 |
|  | MR Egger (accounting for LD correlation) |  | 1.15 | 0.98 | 1.36 | 0.09 |
|  | Weighted median |  | 0.95 | 0.83 | 1.09 | 0.45 |
|  | Inverse variance weighted |  | 1.01 | 0.91 | 1.12 | 0.88 |
|  | Weighted mode |  | 0.94 | 0.80 | 1.12 | 0.53 |
| DACHS all | MR Egger | 6 | 1.25 | 0.52 | 3.02 | 0.64 |
|  | MR Egger (accounting for LD correlation) |  | 0.27 | 0.11 | 0.67 | 4.78x10^-3^ |
|  | Weighted median |  | 1.11 | 0.70 | 1.75 | 0.67 |
|  | Inverse variance weighted |  | 1.14 | 0.77 | 1.68 | 0.53 |
|  | Weighted mode |  | 1.01 | 0.55 | 1.84 | 0.98 |
| DACHS aspirin users | MR Egger | 6 | 0.95 | 0.13 | 6.69 | 0.96 |
|  | MR Egger (accounting for LD correlation) |  | 0.02 | 2.17x10^-3^ | 0.13 | 9.45x10^-5^ |
|  | Weighted median |  | 1.06 | 0.39 | 2.91 | 0.90 |
|  | Inverse variance weighted |  | 1.02 | 0.44 | 2.40 | 0.96 |
|  | Weighted mode |  | 1.14 | 0.32 | 4.03 | 0.84 |
| DACHS aspirin non-users | MR Egger | 6 | 1.26 | 0.44 | 3.61 | 0.68 |
|  | MR Egger (accounting for LD correlation) |  | 0.46 | 0.18 | 1.13 | 0.09 |
|  | Weighted median |  | 1.02 | 0.57 | 1.83 | 0.94 |
|  | Inverse variance weighted |  | 1.26 | 0.78 | 2.01 | 0.35 |
|  | Weighted mode |  | 0.97 | 0.45 | 2.11 | 0.94 |

Abbreviations: OR, odds ratio; LCI, lower confidence interval; UCI, upper confidence interval.

Supplementary Table 5- Results of the Q statistic heterogeneity test for pathway SNPs

| Study | Method | Q statistic | Degrees of freedom | P value |
| --- | --- | --- | --- | --- |
| GECCO | MR Egger | 4.20 | 4 | 0.38 |
|  | IVW | 4.21 | 5 | 0.52 |
| DACHS all | MR Egger | 0.86 | 4 | 0.93 |
|  | IVW | 0.92 | 5 | 0.97 |
| DACHS aspirin user | MR Egger | 0.12 | 4 | 1.00 |
|  | IVW | 0.13 | 5 | 1.00 |
| DACHS aspirin non-user | MR Egger | 2.21 | 4 | 0.70 |
|  | IVW | 2.21 | 5 | 0.82 |

Supplementary Table 6- Genome-wide SNPs used as instruments

| SNP | Effect allele | Other allele | Beta | SE | LCI | UCI | EAF | Direction | Chr | Position | P value | Sample size | LD clumping R^2^ |
| --- | --- | --- | --- | --- | --- | --- | --- | --- | --- | --- | --- | --- | --- |
| rs7498776 | T | C | -0.13 | 0.02 | -0.17 | -0.09 | 0.90 | -- | 16 | 20611149 | 8.70 x10^-11^ | 14149 | 0.001 and 0.8 |
| rs11642648 | A | G | 0.18 | 0.03 | 0.11 | 0.24 | 0.04 | ++ | 16 | 20610955 | 3.58x10^-08^ | 14149 | 0.8 |
| rs7499557 | T | G | -0.25 | 0.04 | -0.34 | -0.17 | 0.98 | -- | 16 | 20557620 | 1.29x10^-09^ | 14149 | 0.8 |
| rs8062555 | C | G | -0.27 | 0.04 | -0.35 | -0.18 | 0.98 | -- | 16 | 20566287 | 2.07 x10^-10^ | 14149 | 0.8 |

Abbreviations: SE, standard error; LCI, lower confidence interval; UCI, upper confidence interval; Chr, chromosome; LD, linkage disequilibrium.

Supplementary Table 7- Genome-wide SNP associations with CRC using the other MR methods

| Study | Method | N SNP | OR | LCI | UCI | P value |
| --- | --- | --- | --- | --- | --- | --- |
| GECCO | MR Egger | 4 | 0.84 | 0.58 | 1.22 | 0.46 |
|  | MR Egger (accounting for LD correlation) | 4 | 1.16 | 0.99 | 1.38 | 0.08 |
|  | Weighted median | 4 | 1.00 | 0.87 | 1.14 | 0.96 |
|  | Inverse variance weighted | 4 | 1.03 | 0.92 | 1.15 | 0.67 |
|  | Weighted mode | 4 | 0.96 | 0.80 | 1.16 | 0.73 |
| DACHS all | MR Egger | 4 | 1.00 | 0.24 | 4.24 | 1.00 |
|  | MR Egger (accounting for LD correlation) | 4 | 0.11 | 0.04 | 0.28 | 4.52x10^-6^ |
|  | Weighted median | 4 | 1.03 | 0.64 | 1.65 | 0.90 |
|  | Inverse variance weighted | 4 | 1.06 | 0.69 | 1.63 | 0.79 |
|  | Weighted mode | 4 | 1.01 | 0.54 | 1.89 | 0.98 |
| DACHS aspirin users | MR Egger | 4 | 1.60 | 0.07 | 38.99 | 0.80 |
|  | MR Egger (accounting for LD correlation) | 4 | 2.47x10^-3^ | 2.37x10^-4^ | 0.03 | 5.07x10^-7^ |
|  | Weighted median | 4 | 0.97 | 0.32 | 2.96 | 0.96 |
|  | Inverse variance weighted | 4 | 0.99 | 0.38 | 2.57 | 0.99 |
|  | Weighted mode | 4 | 0.85 | 0.22 | 3.29 | 0.83 |
| DACHS aspirin non-users | MR Egger | 4 | 0.82 | 0.15 | 4.55 | 0.84 |
|  | MR Egger (accounting for LD correlation) | 4 | 0.23 | 0.10 | 0.52 | 4.15x10^-4^ |
|  | Weighted median | 4 | 1.03 | 0.58 | 1.83 | 0.92 |
|  | Inverse variance weighted | 4 | 1.10 | 0.66 | 1.84 | 0.71 |
|  | Weighted mode | 4 | 0.99 | 0.47 | 2.06 | 0.98 |

Abbreviations: OR, odds ratio; LCI, lower confidence interval; UCI, upper confidence interval.

Supplementary Table 8- Results of the Q statistic heterogeneity test for genome-wide SNPs

| Study | Method | Q statistic | Degrees of freedom | P value |
| --- | --- | --- | --- | --- |
| GECCO | MR Egger | 1.46 | 2 | 0.48 |
|  | IVW | 2.64 | 3 | 0.45 |
| DACHS all | MR Egger | 0.26 | 2 | 0.88 |
|  | IVW | 0.26 | 3 | 0.97 |
| DACHS aspirin users | MR Egger | 0.36 | 2 | 0.84 |
|  | IVW | 0.45 | 3 | 0.93 |
| DACHS aspirin non-users | MR Egger | 0.52 | 2 | 0.77 |
|  | IVW | 0.64 | 3 | 0.89 |
